# An Evaluation of the Vulnerable Physician Workforce in the United States During the Coronavirus Disease-19 Pandemic

**DOI:** 10.1101/2020.03.26.20044263

**Authors:** Rohan Khera, Lovedeep Singh Dhingra, Snigdha Jain, Harlan M Krumholz

**Author notes:** **Correspondence to:** Dr. Rohan Khera; 5323 Harry Hines Blvd, E5.730, Dallas, TX - 75219; @rohan_khera.

## Abstract

**Background:** The coronavirus disease-19 (COVID-19) pandemic threatens to overwhelm the healthcare resources of the country, but also poses a personal hazard to healthcare workers, including physicians.  To address the potential impact of excluding physicians with a high risk of adverse outcomes based on age, we evaluated the current patterns of age of licensed physicians across the United States.

**Methods:** We compiled information from the 2018 database of actively licensed physicians in the Federation of State Medical Boards (FSMB) across the US. Both at a national- and the state-level, we assessed the number and proportion of physicians who would be at an elevated risk due to age over 60 years.

**Results:** Of the 985,026 licensed physicians in the US, 235857 or 23.9% were aged 25-40 years, 447052 or 45.4% are 40-60 years, 191794 or 19.5% were 60-70 years, and 106121 or 10.8% were 70 years or older. Age was not reported in 4202 or 0.4% of physicians. Overall, 297915 or 30.2% of physicians were 60 years of age or older, 246167 (25.0%) 65 years and older, and 106121 (10.8%) 70 years or older. States in the US reported that a median 5470 licensed physicians (interquartile range [IQR], 2394 to 10108) were 60 years of age or older. Notably, states of North Dakota (n=1180) and Vermont (n = 1215) had the lowest and California (n=50786) and New York (n=31582) the highest number of physicians over the age of 60 years (Figure 1). Across states, the median proportion of physicians aged 60 years and older was 28.9% (IQR, 27.2%, 31.4%), and ranged between 25.9% for Nebraska to 32.6% for New Mexico (Figure 2).

**Discussion:** Older physicians represent a large proportion of the US physician workforce, particularly in states with the worst COVID-19 outbreak. Therefore, their exclusion from patient care will be impractical. Optimizing care practices by limiting direct patient contact of physicians vulnerable to adverse outcomes from COVID-19, potentially by expanding their participation in telehealth may be a strategy to protect them.

## Background

The coronavirus disease-19 (COVID-19) pandemic threatens to overwhelm the healthcare resources of the country,^1^ but also poses a personal hazard to healthcare workers, including physicians.^2^ Physicians are at an elevated risk of acquiring the disease through exposure to patients who may be symptomatic with the disease or its asymptomatic carriers across the spectrum of clinical specialties. Notably, the physician workforce is not only at risk of losing time spent in clinical care due to these exposures, but at a personal risk from severe disease that requires hospitalization and is associated with high morbidity and mortality. Notably, physicians 60 years of age and older are at a particularly elevated risk, with 80% of deaths in China concentrated in this age group.^3,4^ In the early experience in the US, nearly half of all hospitalizations and intensive care unit admissions, and nearly 80% of deaths have occurred in this age group as well.^4^ To address the potential impact of excluding physicians with a high risk of adverse outcomes based on age, we evaluated the current patterns of age of licensed physicians across the United States.

## Methods

We used the 2018 database of physicians from Federation of State Medical Boards (FSMB) that includes all actively licensed physicians across the US.^5^ The FSMB maintains a database of all licensed physicians in each of the states and US territories compiled during the licensing process, which are sampled biennially and publicly reported.^5^ We extracted publicly available summary data for the physician age in 5-year age bins, nationally and across each of the states. Both at a national and the state-level, we used descriptive statistics for the age distribution of physicians and assessed the number and proportion of physicians in each state who would be at elevated risk due to age (age >60 years). We used Stata 16 (College Station, TX) for all analyses.

## Results

Of the 985,026 licensed physicians in the US, 235857 or 23.9% were aged 25-40 years, 447052 or 45.4% are 40-60 years, 191794 or 19.5% were 60-70 years, and 106121 or 10.8% were 70 years or older. Age was not reported in 4202 or 0.4% of physicians. Overall, 297915 or 30.2% of physicians were 60 years of age or older, 246167 (25.0%) 65 years and older, and 106121 (10.8%) 70 years or older. States in the US reported that a median of 5470 licensed physicians (interquartile range [IQR], 2394 to 10108) were 60 years of age or older. Notably, states of North Dakota (n=1180) and Vermont (n = 1215) had the lowest and California (n=50786) and New York (n=31582) the highest number of physicians over the age of 60 years (**Figure 1**). Across states, the median proportion of physicians aged 60 years and older was 28.9% (IQR, 27.2%, 31.4%), and ranged between 25.9% for Nebraska to 32.6% for New Mexico (**Figure 2**).

**Figure 1:**
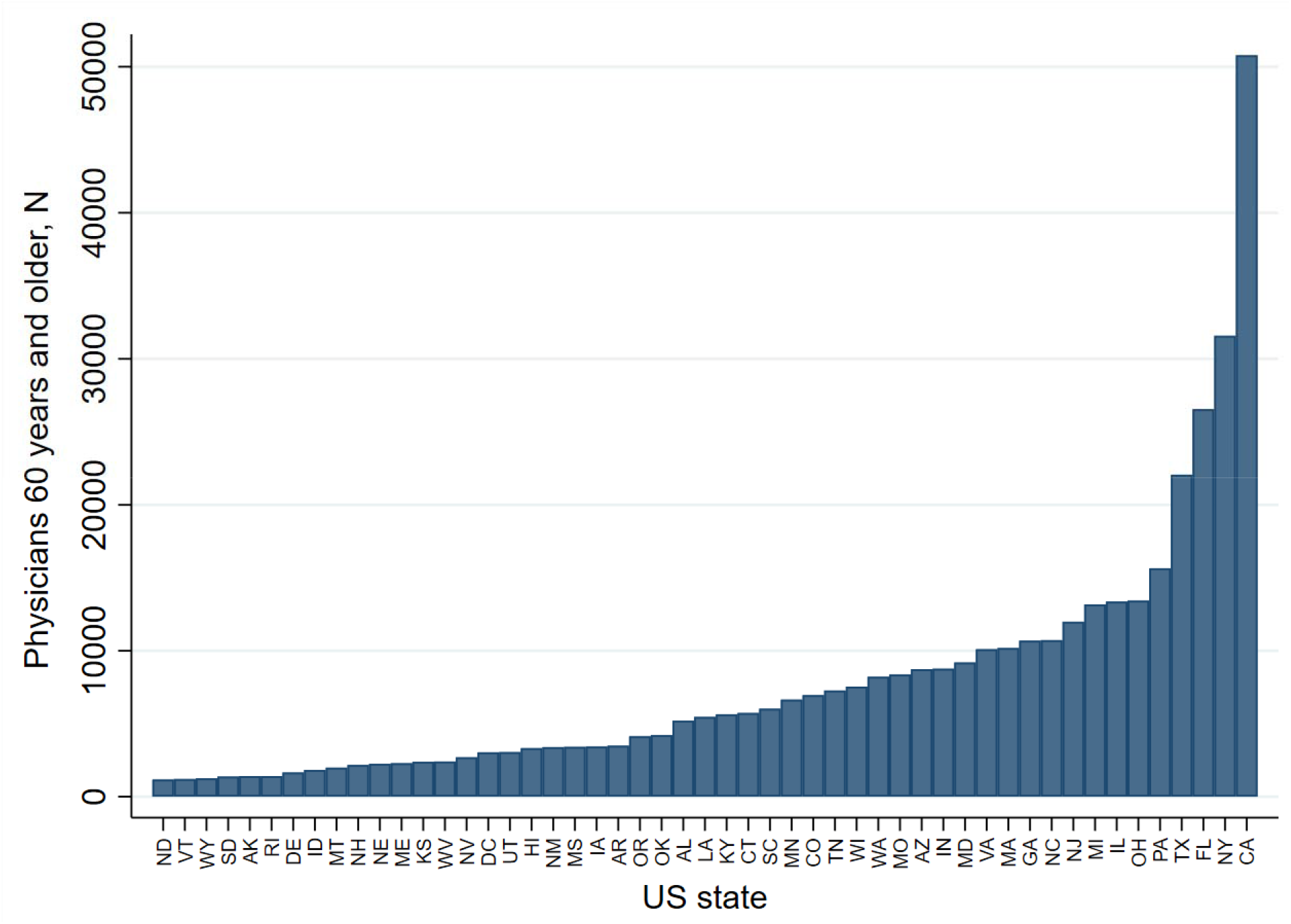
Number of physicians in each state or United States territory 60 years of age and older.

**Figure 2:**
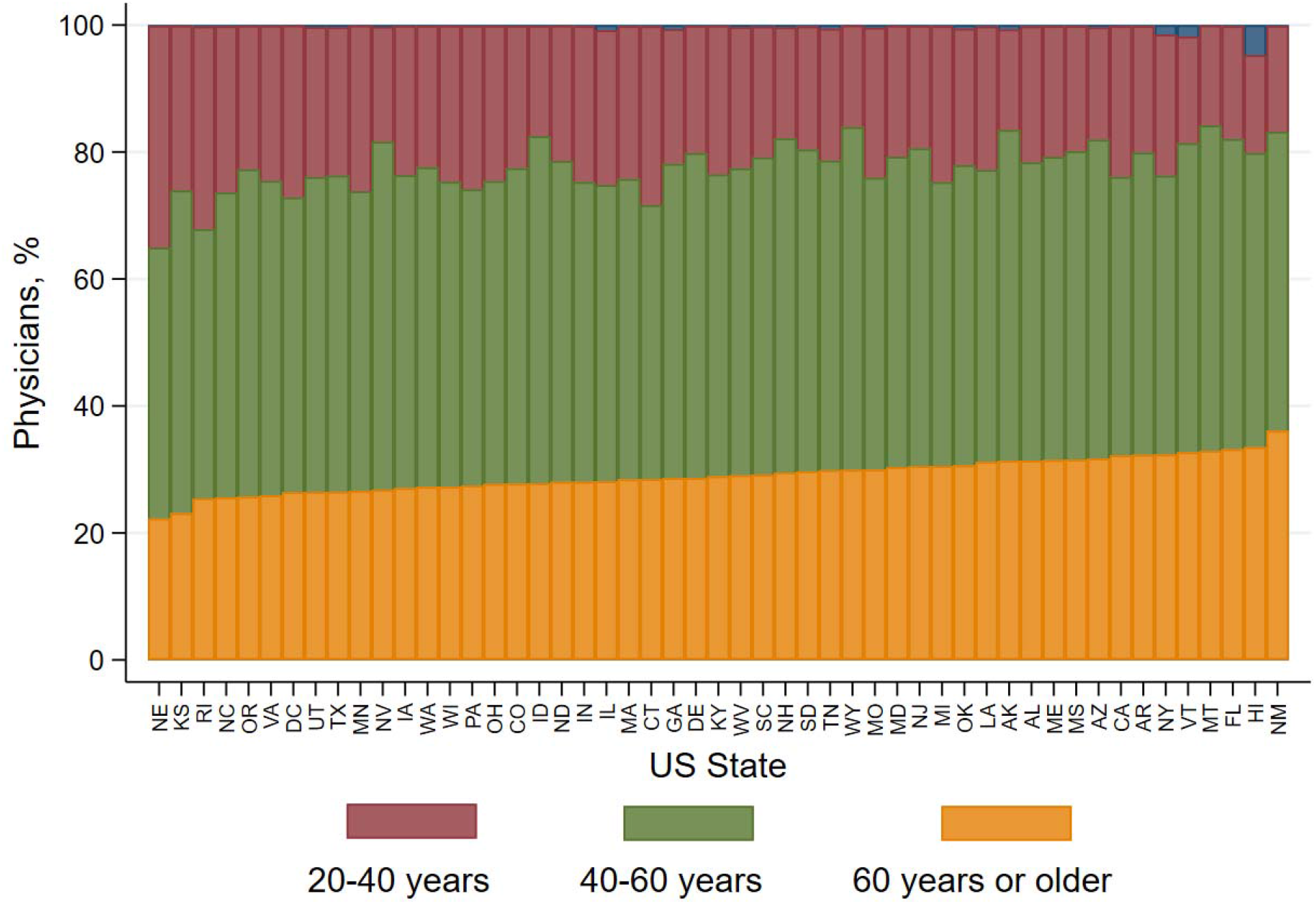
Proportion of physicians in states and United States territories across age groups. The values in blue represent physicians without reported age.

## Discussion

Nearly 1 in 3 licensed physicians in the US and each of the states are over the age of 60 years, representing nearly 300,000 currently licensed physicians in the US. Physicians in the states of California and New York – the two states with the largest outbreaks of COVID-19 also have the highest number of physicians 60 years or older.

The study has certain limitations. We do not have information on the specialty expertise of physicians, as some physicians may be more prone to encountering patients with COVID-19. However, as many individuals in the community may be asymptomatic carriers, physicians across specialties are at risk of acquiring the disease as a part of the patient contact during care delivery. Further, we are unable to identify the health status of physicians that may modify their risk or make some younger physicians vulnerable. Finally, physicians may be licensed in more than one state. National estimates, however, represent the number of unique physicians.

In conclusion, older physicians represent a large proportion of the US physician workforce, particularly in states with the worst COVID-19 outbreak. Therefore, their exclusion from patient care will be impractical. Optimizing care practices by limiting direct patient contact of physicians vulnerable to adverse outcomes from COVID-19, potentially by expanding their participation in telehealth may be a strategy to protect them.

## Data Availability

The data for these analyses are publicly available on the FSMB website.

https://www.fsmb.org/physician-census/

